# Methylation profiling in the Million Veteran Program: design, quality control, and smoking-associated epigenetic signatures

**DOI:** 10.64898/2026.04.22.26351491

**Authors:** Patrick A. Schreiner, Kyriacos Markianos, Michael Francis, Brendan Despard, Bryan R. Gorman, Iskander Said, Frederick Dong, Shruti Gautam, Daniel Dochtermann, Yunling Shi, Poornima Devineni, Colette Kirkpatrick, Nikolay Khazanov, Jennifer Moser, VA Million Veteran Program (MVP), Grant D. Huang, Sumitra Muralidhar, Phillip S. Tsao, Saiju Pyarajan

## Abstract

The Million Veteran Program (MVP) represents the largest and one of the most diverse single cohorts associated with longitudinal Electronic Health Record data (EHR) data. We profiled a subset of samples from MVP using the Illumina Infinium MethylationEPIC Beadchip (EPIC array) to generate one of the largest single cohort methylation dataset to-date. Methylation profiles were analyzed for 45,460 total individuals, with the most populous ancestries composed of 27,455 Europeans, 11,798 African Americans, and 4,859 Admixed Americans. We detail the strict quality control standards implemented to ensure the most robust method of methylation profiling of the MVP cohort. This dataset was then applied to evaluate the effects of smoking exposure on DNA methylation in MVP participants. Ancestry-stratified epigenome-wide association studies (EWAS) of smoking status (ever/never) were performed using over 750,000 probes with certifiable signal. Our multi-ancestry meta-analysis demonstrates replicability with existing EWAS and identifies 3,207 novel probe-smoking associations unlocked via the depth and breadth of data in this cohort.

## Introduction

DNA methylation is the central epigenetic mechanism through which environmental exposures can induce changes in gene regulation throughout life^1–5^. Biobank-scale cohorts have proven effective in validating canonical DNA methylation associations, identifying novel biomarkers, and providing insight into phenotypic mechanisms^6–13^. However, expanding the scale, ancestral diversity, and linkage to genetic and phenotypic data of modern methylation cohorts remains essential to enabling high-resolution discovery of associations between methylation signatures and disease.

Cigarette smoking, one of the leading contributing factors of preventable death worldwide^14–17^, leads to dysregulation of epigenetic programming, the consequences of which are observable in blood methylation patterns. There is increasing evidence that adverse health outcomes associated with smoking, such as cardiovascular disease, inflammation, and cancer, may be partly driven by epigenetic modifications^18–20^, making this an important area of investigation. Previous smoking epigenome-wide association studies (EWAS) performed using the Infinium HumanMethylation450 BeadChip^21^ uncovered many probes found to be differentially methylated in smokers, including highly significant associations at *AHRR, F2RL3* and *RARA*^22–25^. The Illumina Infinium MethylationEPIC array (EPICv1)^26^, which supersedes the 450K platform, interrogates >850,000 CpG sites across the genome, including enhancer regions and other regulatory loci not covered on the 450K chip^27^. This expanded coverage has enabled the discovery of thousands of additional smoking-associated CpGs^28–36^. Overall, these findings have unlocked insight into the broad and complex effects of smoking exposure on an individual’s epigenetic landscape.

Most large EWAS of smoking exposure to-date have been conducted in predominately European cohorts. However, studies using African and Hispanic participants have replicated many of the top smoking-associated CpGs^23,34,37^. The convergence of evidence from diverse cohorts has solidified the most robust methylation associations with smoking but has also indicated population-specific differences^23,36,38,39^, underscoring the importance of multi-ancestry replication and consideration of population-specific contexts.

Here, we present the Million Veteran Program’s (MVP) methylation cohort, which represents the largest, and most diverse, single cohort of EPICv1 array samples to date (n=45,460). Our study describes the quality control of data generated by this array and provides novel insights into how cigarette smoking alters the epigenome via EWAS in MVP participants of European (EUR), African (AFR) and Admixed American (AMR) ancestry.

## Results

### MVP Methylation Cohort

Blood collected from veteran volunteers from sites across the United States was shipped overnight on blue ice to the central biorepository, where buffy coat and plasma were separated and stored at −80^0^C. DNA was extracted from buffy coat and sent for profiling using the EPICv1 methylation array^26^ (Figure 1A). The MVP cohort profiles 27,455 EUR, 11,798 AFR, 4,859 AMR, 608 East Asian (EAS), 43 South Asian (SAS), and 697 individuals of indeterminate genetically inferred ancestry (Figure 1B; Supplementary Data 1). Individuals in this study were predominantly male, of advanced age, and exhibited an elevated level of exposure to smoking relative to other cohorts (Figure 1C)^40^. A subset of the methylation cohort was consortium-selected, which overrepresented specific phenotypes relative to random profiling in the MVP cohort, including PTSD, chronic kidney, peripheral artery, and cardiovascular diseases (Figure 1D).

**Figure 1.**
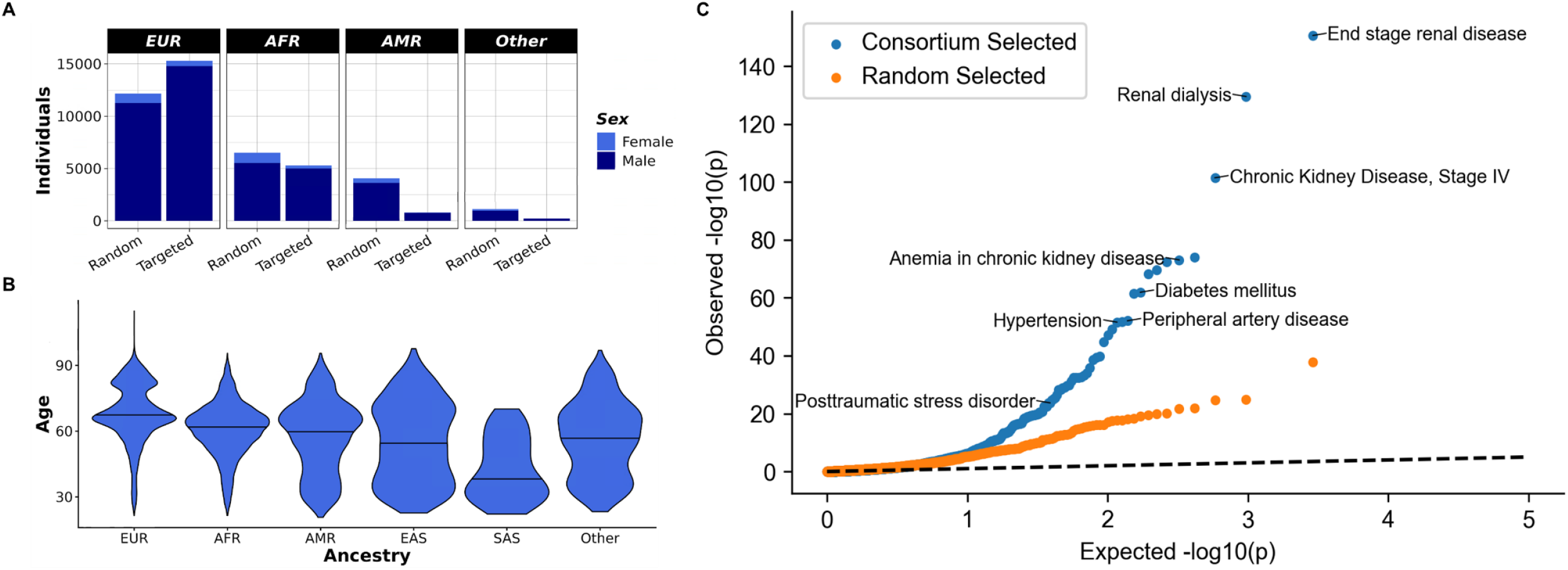
MVP Methylation Cohort. (A) The distribution of MVP participants’ sex and description of inclusion for profiled individuals profiled categorized by ancestry. (B) Age distributions of profiled individuals by GIA. The median age per ancestry is designated with a line. (C) Q-Q plot demonstrating the targeted nature of phenotypes in the methylation cohort relative to the full MVP cohort.

### Data Quality Control

We developed a standardized workflow to assess probe and sample inclusion in our study (Figure 2A). Probes that have been previously flagged for potential artifact production were excluded (see Methods)^41^. Probe retention required a consistent, certifiable signal as defined by *p*-value with out-of-band array hybridization (pOOBAH)s (Figure 2B; see Methods)^42^. After exclusions based upon probe design and signal quality, 752,930 high-quality, autosomal probes were retained for EWAS analysis.

**Figure 2.**
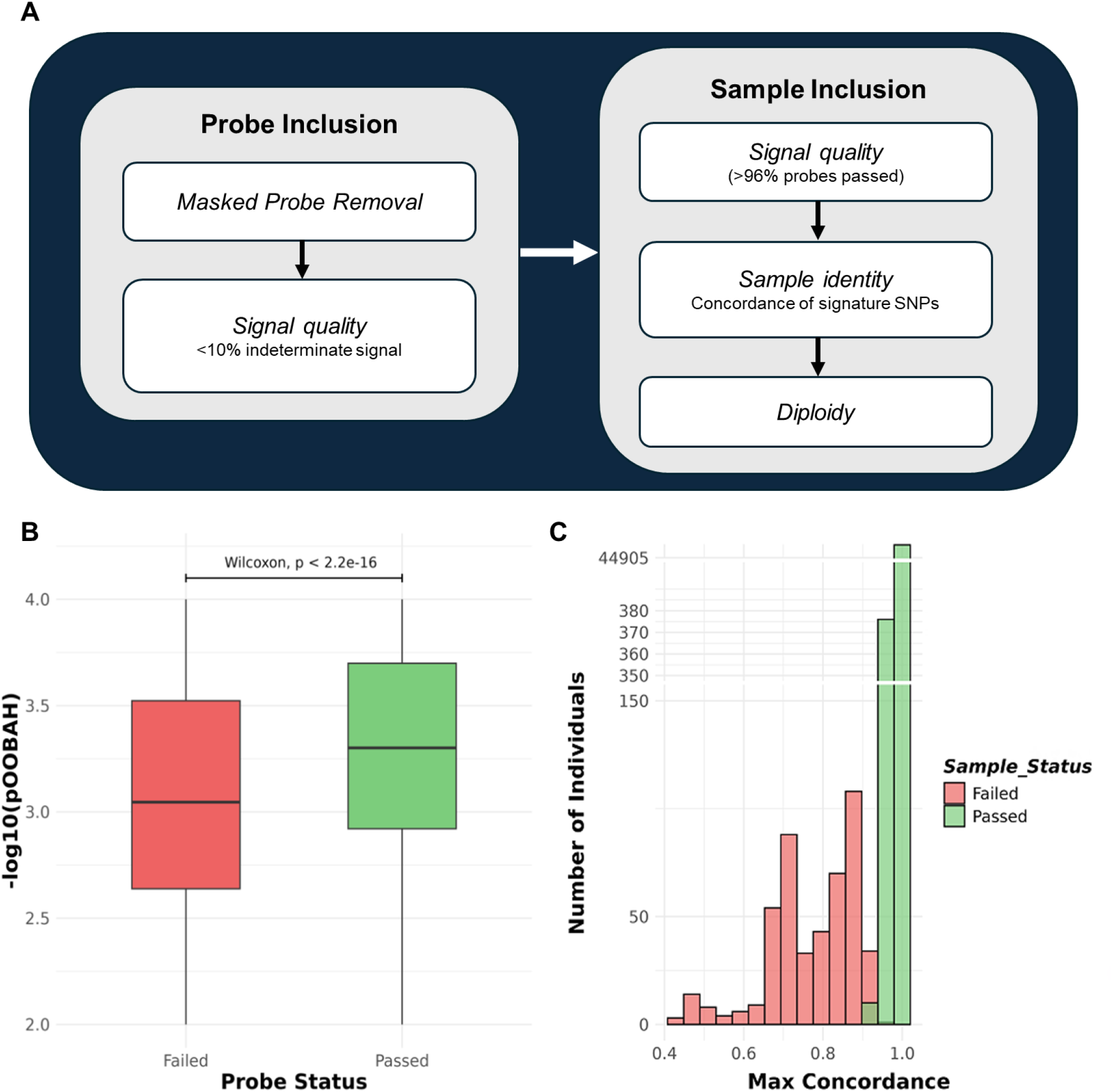
Data Quality Control and Sample Identity Confirmation. (A) Workflow for probe and sample exclusion. (B) Probe pOOBAH values indicating signal assurance for probes that were included and excluded from analysis (C) Signature SNP concordance analysis for individuals that were profiled using both genotyping and methylation arrays. Samples that passed SNP fingerprinting are shown in green and samples excluded at this step are shown in red.

Samples were then evaluated for successful methylation profiling by identifying certifiable signal using pOOBAH (Figure 2A; Methods). Individual sample identity was confirmed using existing SNP-chip data. We required concordance between SNP-chip and methylation array derived genotypes (Figure 2C; Methods)^42,43^. Only samples with expected sex diploidy were retained^43^.

### High-resolution EWAS of smoking status

We performed EWAS of binary smoking status (ever/never) in MVP EUR, AFR, and AMR using meffil^44^ (Supplementary Data 2; Supplementary Fig. 1). Raw EWAS *p*-value output showed signs of systematic inflation (*λ*>2 in full ancestry-stratified cohorts), therefore we used bacon^45^ to generate corrected test statistics. The bacon-corrected *p*-values were within a well-controlled range for EWAS (*λ*=1.03 to 1.43; Supplementary Data 3). Because the full methylation cohort contained enrichment for certain phenotypes, we ran EWAS using only the randomly selected samples within each ancestry as a sensitivity analysis for potential biases; sample characteristics such as age, sex, and BMI were similar between the full and randomly selected cohorts (Supplementary Data 2). We observed high epigenome-wide correlation of *Z*-scores between the full and randomly selected cohorts for EUR, AFR, and AMR ancestries (*ρ*=0.780, 0.828, and 0.912, respectively; Supplementary Data 3), therefore we focus henceforth on the more highly powered full cohorts.

Significance was defined by bacon-corrected *p*-values reaching a Bonferroni-corrected epigenome-wide threshold (α=0.05/752,930 probes). In MVP, we identified 8,453, 5,411, and 182 significant EWAS probes in EUR, AFR, and AMR, respectively (Supplementary Data 3). Across MVP ancestries we observed a high proportion of overlapping significant probes; for example, 78% of AFR probes (4,216 of 5,412) were significant in EUR, and all 182 significant AMR probes were also significant in both EUR and AFR (Supplementary Data 4, 5). Top hits were consistent across ancestries, including known smoking probes at *AHRR, F2RL3, RARA, PRSS23*, and *ALPPL2* (Supplementary Fig. 1). Among probes that were significant in one or more MVP ancestries, their *Z*-scores were highly correlated, with pairwise *ρ* ranging from 0.852-0.951 (Supplementary Data 5). Across 9,651 probes that were significant in one or more ancestry, 9,401 (97.4%) had the same effect direction in EUR, AFR, and AMR.

Additionally, we compared our primary (“forward”) regression model in MVP EUR, where methylation M-values were modeled as the outcome and ever smoking as the predictor, to a “reverse” model that swapped these roles. The Bacon-corrected −log_10_(*p*) values were nearly identical between model directions (*r*=0.996), indicating that the strength of association signals was not sensitive to this choice (Supplementary Fig. 2). The “reverse” models were run as a sensitivity analysis and are not used in subsequent results or interpretation; unless otherwise noted, all analyses refer to the “forward” models.

We performed replication of our results using an EWAS meta-analysis of current smoking in ≤15,014 European adults by Hoang et al.^28^. We identified 54%, 64%, and 99% of our significant MVP probes in Hoang et al.^28^ for EUR, AFR, and AMR, respectively, with upwards of 99.9% of shared significant probes having the same effect direction (Supplementary Data 3). Among probes that were significant in one or more cohort between MVP and Hoang et al.^28^, pairwise *ρ* ranged from 0.781 (EUR) to 0.828 (AMR). (Supplementary Data 4, 5). To perform a direct comparison of effect sizes between MVP EUR and Hoang et al.^28^, we generated EWAS summary statistics for MVP EUR using methylation beta values (as opposed to M-values). We observed a strong correlation of effect sizes across the 14,617 probes that were significant in one or both EWAS (*r*=0.892; Supplementary Fig. 3).

We then performed a meta-analysis of MVP EUR, AFR, and AMR EWAS results, yielding a combined multi-ancestry cohort of 40,756 MVP samples. Meta-analysis identified 19,067 significant smoking probes (Table 1; Supplementary Data 6). Although most probes were replicated across ancestries, many exhibited cross-ancestry heterogeneity in effect estimates. We observed a strong correlation of effect sizes between the MVP meta-analysis and Hoang et al.^28^ among probes that were significant in one or both cohorts (*ρ*= 0.787; Supplementary Data 5).

**Table 1.** Top 25 CpGs differentially methylated in relation to ever smoking. Effects correspond to the MVP multi-ancestry fixed effects meta-analysis (N=40,756). Effect, standard error (SE) and *p*-value shown are post-bacon-correction.

| CpG | CHR:POS<br>(hg19) | Nearest<br>gene | Effect | SE | *p*-value | Direction<br>(EUR, AFR, AMR) |
| --- | --- | --- | --- | --- | --- | --- |
| cg05575921 | 5:373,378 | AHRR | -0.974 | 0.013 | $5.69 \times 10^{-1106}$ | -, -, - |
| cg14391737 | 11:86,513,429 | PRSS23 | -0.383 | 0.006 | $1.03 \times 10^{-849}$ | -, -, - |
| cg21566642 | 2:233,284,661 | ALPPL2 | -0.384 | 0.006 | $2.00 \times 10^{-817}$ | -, -, - |
| cg01940273 | 2:233,284,934 | ALPPL2 | -0.234 | 0.004 | $9.08 \times 10^{-627}$ | -, -, - |
| cg03636183 | 19:17,000,585 | F2RL3 | -0.270 | 0.005 | $1.07 \times 10^{-545}$ | -, -, - |
| cg00475490 | 11:86,517,110 | PRSS23 | -0.338 | 0.006 | $3.74 \times 10^{-519}$ | -, -, - |
| cg21911711 | 19:16,998,668 | F2RL3 | -0.282 | 0.006 | $2.00 \times 10^{-489}$ | -, -, - |
| cg17739917 | 17:38,477,572 | RARA | -0.236 | 0.005 | $7.05 \times 10^{-462}$ | -, -, - |
| cg06644428 | 2:233,284,112 | ALPPL2 | -0.469 | 0.010 | $6.61 \times 10^{-454}$ | -, -, - |
| cg23576855 | 5:373,299 | AHRR | -0.524 | 0.012 | $6.83 \times 10^{-434}$ | -, -, - |
| cg21161138 | 5:399,360 | AHRR | -0.212 | 0.005 | $2.07 \times 10^{-413}$ | -, -, - |
| cg18110140 | 15:75,350,380 | MEPCE | -0.194 | 0.005 | $1.71 \times 10^{-385}$ | -, -, - |
| cg19859270 | 3:98,251,294 | GPR15 | -0.206 | 0.005 | $5.89 \times 10^{-367}$ | -, -, - |
| cg14466441 | 6:11,392,193 | SMAP1 | -0.190 | 0.005 | $2.50 \times 10^{-360}$ | -, -, - |
| cg10711136 | 11:86,515,053 | PRSS23 | -0.214 | 0.005 | $3.62 \times 10^{-344}$ | -, -, - |
| cg09935388 | 1:92,947,588 | GF1 | -0.375 | 0.010 | $8.81 \times 10^{-333}$ | -, -, - |
| cg17087741 | 2:233,283,010 | ALPPL2 | -0.249 | 0.006 | $2.95 \times 10^{-323}$ | -, -, - |
| cg11660018 | 11:86,510,915 | PRSS23 | -0.138 | 0.004 | $4.79 \times 10^{-323}$ | -, -, - |
| cg19572487 | 17:38,476,024 | RARA | -0.167 | 0.004 | $8.75 \times 10^{-322}$ | -, -, - |
| cg14753356 | 6:30,720,108 | HLA-DPA1 | -0.142 | 0.004 | $1.09 \times 10^{-310}$ | -, -, - |
| cg24859433 | 6:30,720,203 | HLA-DPA1 | -0.221 | 0.006 | $3.58 \times 10^{-304}$ | -, -, - |
| cg15342087 | 6:30,720,209 | HLA-DPA1 | -0.221 | 0.006 | $7.80 \times 10^{-297}$ | -, -, - |
| cg03329539 | 2:233,283,329 | ALPPL2 | -0.122 | 0.003 | $7.91 \times 10^{-294}$ | -, -, - |
| cg00045592 | 1:160,714,299 | SLAMF7 | -0.155 | 0.004 | $6.58 \times 10^{-290}$ | -, -, - |
| cg25189904 | 1:68,299,493 | GNG12 | -0.242 | 0.007 | $9.16 \times 10^{-282}$ | -, -, - |

Consistent with expectations, cg05575921 in *AHRR* was the most significant probe and had a strong negative effect in all MVP EWAS cohorts (effect_MVPmeta_=-0.974; standard error (se)=0.013; *P*_MVPmeta_=5.69×10^−1106^; Table 1). Novelty of our findings was assessed against all hits found in previously published smoking EWAS (32 publications; Supplementary Data 7). We identified a total of 3,207 novel smoking EWAS probes, including 3,071 present in the multi-ancestry meta-analysis (Table 2; Supplementary Data 3).

**Table 2.** Top 25 novel CpGs differentially methylated in relation to ever smoking. Novel probes were not reported as significant in any previous smoking EWAS. Effects correspond to the MVP multi-ancestry fixed effects meta-analysis (N= 40,756). Effect, standard error (SE) and *p*-value shown are post-bacon-correction

| CpG | CHR:POS (hg19) | Nearest gene | Effect | SE | p-value | Direction<br>(EUR, AFR, AMR) |
| --- | --- | --- | --- | --- | --- | --- |
| cg03909289 | 19:30,206,815 | *C19orf12* | -0.054 | 0.004 | $1.93 \times 10^{-37}$ | -, -, - |
| cg21734651 | 10:75,905,043 | *AP3M1* | -0.082 | 0.006 | $7.16 \times 10^{-37}$ | -, -, - |
| cg22587479 | 2:219,738,226 | *WNT6* | 0.129 | 0.010 | $7.89 \times 10^{-36}$ | +, +, + |
| cg27052582 | 20:37,062,821 | *SNORA71D* | -0.057 | 0.005 | $2.20 \times 10^{-33}$ | -, -, - |
| cg23716779 | 19:46,998,790 | *PNMAL2* | 0.057 | 0.005 | $2.51 \times 10^{-33}$ | +, +, + |
| cg11414254 | 5:176,449,012 | *ZNF346* | -0.053 | 0.005 | $6.61 \times 10^{-32}$ | -, -, - |
| cg14620941 | 9:139,776,929 | *TRAF2* | -0.050 | 0.004 | $2.05 \times 10^{-31}$ | -, -, - |
| cg08228578 | 12:57,624,193 | *SHMT2* | -0.025 | 0.002 | $1.91 \times 10^{-27}$ | -, -, - |
| cg27394111 | 10:26,612,372 | *GAD2* | 0.045 | 0.004 | $1.79 \times 10^{-26}$ | +, +, + |
| cg19168482 | 6:116,576,498 | *TSPYL4* | -0.063 | 0.006 | $4.36 \times 10^{-26}$ | -, -, - |
| cg19799178 | 6:37,136,661 | *PIM1* | -0.037 | 0.004 | $4.61 \times 10^{-26}$ | -, -, - |
| cg14271520 | 5:123,149,749 | *CSNK1G3* | -0.072 | 0.007 | $4.67 \times 10^{-26}$ | -, -, - |
| cg10604476 | 19:10,403,908 | *ICAM5* | 0.122 | 0.012 | $2.83 \times 10^{-25}$ | +, +, + |
| cg02974977 | 1:67,893,397 | *SERBP1* | -0.043 | 0.004 | $9.12 \times 10^{-25}$ | -, -, - |
| cg18747276 | 22:29,193,159 | *XBP1* | -0.058 | 0.006 | $1.07 \times 10^{-24}$ | -, -, - |
| cg13766434 | 6:17,981,180 | *KIF13A* | -0.058 | 0.006 | $3.67 \times 10^{-24}$ | -, -, - |
| cg11031221 | 17:70,417,517 | *LINC00673* | -0.044 | 0.004 | $4.69 \times 10^{-24}$ | -, -, - |
| cg10731236 | 3:95,986,723 | *EPHA6* | 0.068 | 0.007 | $4.84 \times 10^{-24}$ | +, +, + |
| cg24516256 | 9:136,213,816 | *MED22* | -0.045 | 0.005 | $3.63 \times 10^{-23}$ | -, -, - |
| cg21577590 | 6:116,576,752 | *TSPYL4* | -0.058 | 0.006 | $6.22 \times 10^{-23}$ | -, -, - |
| cg21482921 | 10:5,084,043 | *AKR1C3* | 0.051 | 0.005 | $8.59 \times 10^{-23}$ | +, +, + |
| cg23587474 | 15:69,970,066 | *PCAT29* | 0.037 | 0.004 | $9.29 \times 10^{-23}$ | +, +, + |
| cg16155661 | 16:30,440,676 | *DCTPP1* | -0.041 | 0.004 | $9.48 \times 10^{-23}$ | -, -, - |
| cg02419562 | 8:8,746,748 | *MFHAS1* | -0.063 | 0.006 | $2.38 \times 10^{-22}$ | -, -, - |
| cg00688546 | 8:48,874,244 | *MCM4* | -0.032 | 0.003 | $4.26 \times 10^{-22}$ | -, -, - |

## Discussion

Here we explore the effect of smoking on individuals in the largest and most genetically diverse cohort of methylation array data currently available. Standardized array-based DNA methylation profiling has enabled large-scale EWAS of environmental exposures, with cigarette smoking being one of the most extensively characterized.

The unique size and diverse ancestral composition of our cohort, paired with the expansive CpG coverage of the EPICv1 array, enabled this study of smoking–methylation associations at unprecedented resolution. Using a conservative genome-wide significance threshold, we identified 3,207 novel associations, substantially extending the known methylation signature of smoking exposure. It is also important to note that EWAS results can capture aspects of biology that are not fully explained by germline genetic variation identified in GWAS^46^, highlighting the complementary nature of these approaches.

Since these data were generated as part of a consortium effort, there is a degree of phenotype selection within the cohort (Figure 1D). However, results from the full cohort demonstrated robustness of smoking epigenetic signature to this phenotype selection when compared to the randomly selected subgroup of individuals (Supplementary Data 3). We caution that robustness to phenotype selection does not necessary imply that EWAS results are insensitive to selection for scans other than smoking status. We encourage researchers to check for sensitivity to selection using approaches similar to the analysis presented here. We further validated our findings by confirming strong effects at canonical smoking-associated probes and by demonstrating consistency with published EWAS of smoking exposure.

Taken together, our study refines and extends characterization of the smoking-associated methylation signature using a uniquely large and diverse cohort. Our results provide high-resolution insight into methylation probes exhibiting both known and novel associations with smoking exposure. These findings enable secondary analyses and meta-analyses that can prioritize epigenetic biomarkers and molecular pathways contributing to disease risk related to smoking exposure, and they provide a resource for future integrative and mechanistic studies.

## Methods

### Ethics/study approval

The VA Central Institutional Review Board (IRB) approved the study protocol. All participants provided informed consent, and all studies conducted at participating centers received approval from IRBs, in accordance with the Declaration of Helsinki.

### Sample Preparation

Genomic DNA was isolated and purified from whole blood using the GenFind V3 kit (Beckman Coulter Cat# CA76409-761). Bisulfite conversion was performed using the ZYMO EZ-96 DNA Methylation^™^Kit (Zymo Research Cat# D5004) in alignment with the Illumina recommendation. DNA was then PCR amplified before profiling on the Infinium MethylationEPIC v1.0 array^26^ (Figure 1A).

### Raw Signal Preprocessing

Raw red/green fluorescence intensity-based data files (IDATs) were processed with SeSAMe (R package v1.8.2). All steps were executed on per-sample SeSAMe SigDF/sset objects, with probe-level detection, background correction, dye-bias correction, and beta value estimation performed within SeSAMe. Signal preprocessing used SeSAMe’s recommended “ind” pipeline. InferTypeIChannel(sset) was initially used to infer/repair Type-I probe channel assignments (including SNP-induced channel switches). Out-of-band background subtraction was performed using the noob(sset). Type-I red/green midpoint intensity normalization was performed using DyeBiasCorrTypeINorm(sset). Following “ind’ processing, we computed DNAm beta values using getBetas()^42^.

### Sample Fingerprinting

All samples described herein were also genotyped using the Thermofisher MVP-1 Axiom array genotypes^43^. Sample identity was verified by assessing genotype concordance between EPICv1 array and MVP-1 genotypes.

For fingerprinting, we used both SNP probes included in the EPICv1 array and imputed SNP genotypes derived from Type I methylation probes. We selected fingerprinting probes using the following criteria: overlap between methylation probe and high-quality, typed or imputed, Affymetrix genotypes, allele frequency > 0.5%, biallelic variants, allele frequency concordance for Methylation and chip-derived genotypes. Low-quality genotypes (Genotyping score ≤ 20)^42^ were set to missing, and comparison was restricted to pairs passing QC for both assays. Overall, 211 variants were chosen for fingerprinting, 30 SNPs, and 181 imputed variants. We computed pairwise genotype concordance:

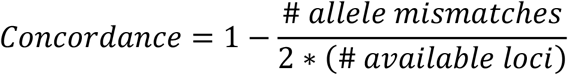

Fingerprinting was carried out in two steps: (1) successful unique mapping of the sample to an Affymetrix genotype assay (SampleMapPass), (2) SNP concordance between the putative sample ID derived from the chip and the sample ID associated with the Methylation assay. For unique mapping, we required (a) high concordance with at least one genotype assay (maxConcordance > 0.95) and (b) ability to discriminate multiple mappings (top2ConcordanceDiff > 0.05). Both thresholds correspond to the empirical false positive rate <10^-3^. We derived the empirical false positive rate for unique mapping using a comparable size random set of genotyped samples that were not sent for methylation; 12 samples satisfied both mapping criteria (0.027%). Only 369 samples failed to satisfy both unique mapping criteria – maxConcordance > 0.95 and top2ConcordanceDiff > 0.05 – during the assessment of methylation-based SNP fingerprinting against the entire genotyped sample set. Among the remaining samples, another 283 failed sample ID matching. These 652 samples were removed from analysis.

### Probe-level quality control

Probe metadata was provided by the Infinium MethylationEPIC v1.0 B5 Manifest File. The nearest gene was assigned to probes that did not have an assignment. We adhered to the recommended, general probe mask to remove DNA methylation array probes that theoretically could produce technically unreliable measurements^41^. The general mask excluded probes that did not map to the human genome, mapped with low confidence (alignment score <35 for either Infinium-I probe, or <10 for mapped but non-unique probes), or, for Infinium-I assays, had discordant mapping of major vs minor alleles. We also excluded probes whose titration correlation <0.9 since probes should show very high correlation with the titrated methylation fraction. We considered SNP-based masks that identify probes whose measurements may be confounded by common genetic variation at or near the single-base extension site. We flagged probes with at least one common SNP (minor allele frequency ≥5%) within 5 bp of the 3′ extension site since these variants can affect probe binding and extension efficiency. Any probe flagged by one or more of these mapping, performance, or SNP-based masks was excluded prior to downstream analyses.

Probe detection was then evaluated using SeSAMe’s pOOBAH^42^ statistic to estimate the probability that a signal is distinguishable from background defined by out-of-band control probes. Probes with certifiable signal, using pOOBAH at a significance threshold of 0.05, were considered present in a given sample. We removed probes that failed pOOBAH signal assurance in ≥ 10% of samples. These filtering criteria reduced the manufacturer’s 866,836 EPIC v1 probes to a high-confidence set of 752,930 used for downstream analysis.

### Sample-level quality control

Sample filtering proceeded as follows: (i) removal of assay controls; (ii) retention of samples with a probe call rate ≥96% based on pOOBAH pass/fail; (iii) quarantine of samples failing identity verification (see below); and (iv) removal of intentional duplicates. After these steps, 45,460 unique biological samples remained for analysis.

Genetically inferred ancestry definition was assigned as previously reported^47^. Smoking status (current, former, never) was ascertained from smoking-related health factors and ICD-9/10 codes for tobacco dependence, using an algorithm previously validated in the VA^48^. Individuals without smoking data were excluded from the EWAS of smoking exposure. Males with sex chromosome aneuploidy, specifically Klienfelter Syndrome (XXY), were identified and removed from the analysis; this resulted in 32 (0.12%), 7 (0.06%), and 3 (0.07%), samples removed from EUR, AFR, and AMR cohorts, respectively.

We performed phenome-wide association scans (PheWAS) to assess which phenotypes were enriched in the consortium-selected and randomly selected subsets compared to the overall MVP cohort. For each subset, we constructed a predictor contrasting samples in the subset with the remaining samples in the most recent MVP genotype data release and tested it for association with 1,456 phenotypes in a Firth logistic regression model, adjusting for age, sex, and genetic ancestry label. Phenotypes were constructed using the PheWAS method; at least two phecode counts on independent days were required for cases, and controls had zero counts.

### Cell Type Deconvolution

To account for blood cell heterogeneity, we estimated cell-type proportions from beta values using meffil.estimate.cell.counts.from.betas()^44^. We used the Houseman deconvolution method to calculate proportion estimates for B cells, CD4+ T cells, CD8+ T cells, eosinophils, monocytes, neutrophils, and NK cells using GSE35069 as a whole-blood data^44,49,50^. Estimated proportions were carried forward as covariates in our EWAS of smoking exposure.

### Epigenome-Wide Association Testing

Ancestry-stratified EWAS of smoking exposure for EUR, AFR, and AMR individuals were calculated using QC-passed probes with the meffil.ewas() function from meffil (R package v1.3.4)^44^. M-values were used because their statistical properties are best suited for differential methylation analysis^51^. We analyzed associations using both a robust linear model using methylation as the outcome, and using a logistic regression with methylation as the exposure, adjusting for cell-type proportion estimates, age, age-squared, smoking status (ever/never), duration of samples storage, first 10 genetic PCs, and Array Scanner ID as follows (Figure 3).

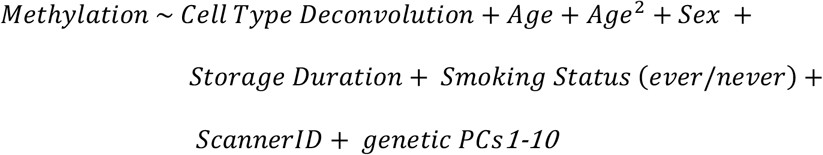

**Figure 3.**
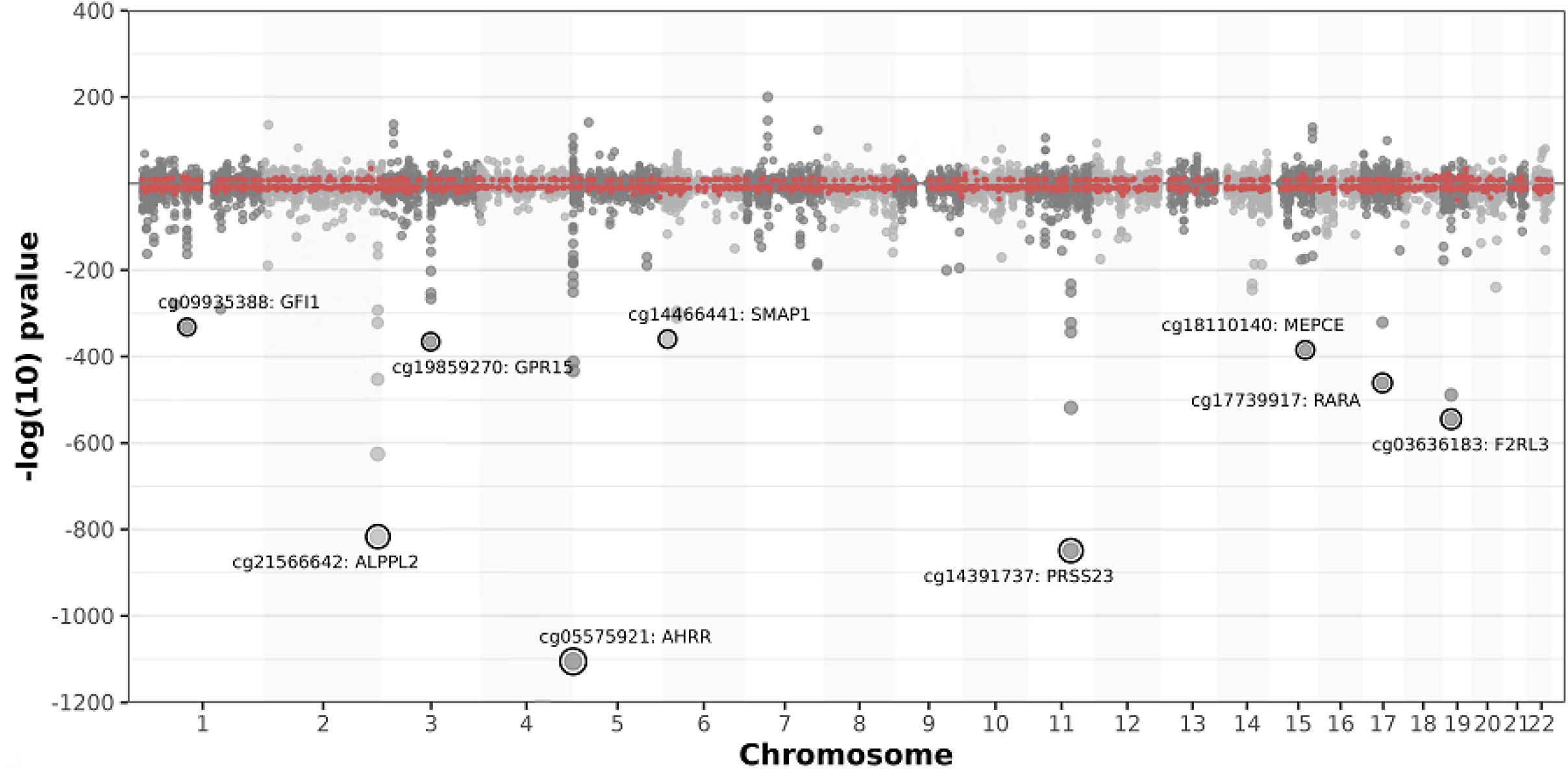
Multi-ancestry EWAS meta-analysis of smoking status in Million Veteran Program. Plots show association *p*-values for ever/never smoking obtained from a two-sided test of the z-statistic. Values below zero reflect sign-coding of −log_10_(*p*-value) by a negative effect estimate. Top gene associations are annotated.

Initial EWAS results resulted in genomic inflation (λ) > 1.2 across all cohorts, suggestive of test statistic bias and inflation. Therefore, inflation and bias reduction was applied using bacon (R package v1.31.1)^45^. Bacon correction was applied to Hoang et al. *p*-values in our summary statistics for comparison purposes^28^ (Supplementary Data 3). Spearman correlation was used for cross-cohort correlation because MVP uses M-values as dependent variable and Hoang et al. (2024) used β-values; except where MVP EUR β-values were generated to directly compare effect sizes in Supplementary Fig. 2, where Pearson’s *r* was used.

The multi-ancestry meta-analysis was calculated using metafor (R package, v4.4.0)^52^. Significance was defined by bacon-corrected *p*-values reaching a Bonferroni-corrected epigenome-wide threshold (α=0.05/752,930 probes). Known vs novel probe association with smoking exposure was defined using all significant hits reported in previously published EWAS (Supplementary Data 6).

### Sensitivity Analyses

We performed a series of sensitivity analyses to ensure robust results. All ancestry-specific EWAS and meta-analysis of smoking exposure were calculated independently using both the full cohort of available individuals and cohorts strictly consisting of randomly-selected individuals. The random-only and full cohort ancestry meta-analyses were calculated independently using both fixed-effect and random-effect models. In MVP EUR, in addition to the primary (‘forward’) models regressing methylation M-values on ever/never smoking status, we fit ‘reverse’ models that swapped the outcome and predictor (ever/never smoking status regressed on methylation M-values) as a check for model robustness (Supplementary Fig. 2).

## Data Availability

Summary data from the present study is available upon reasonable request to the authors

## Contributions

Drafted the manuscript: P.A.S., M.F., K.M., B.D., and S.P. Acquired the data: K.M., Y.S., P.D., C.K., A.K.S., U.S., J.M., G.D.H., S.M., P.S.T., VA MVP, and S.P. Analyzed the data: P.A.S., M.F., B.D., S.G., B.R.G., F.D., D.D., S.S., N.W., K.M, and S.P. Critically revised the manuscript for important intellectual content: M.F., P.A.S., B.R.G., K.M., I.S., D.D., Y.S., and S.P.

## Acknowledgements

This research is based on data from the Million Veteran Program, Office of Research and Development, Veterans Health Administration, and was supported by award #MVP000. This publication does not represent the views of the Department of Veterans Affairs or the United States Government.

